# A prospective study to determine the safety of the geko^®^ neuromuscular electrostimulation device in a pacemaker population

**DOI:** 10.1101/2025.05.20.25327988

**Authors:** Z. Yousef, C. S. Barr

**Author notes:** Trial registration: ClinicalTrials.gov Identifier NCT04391257. Ethics: UK NHS REC REF: 20/WA/0166 (IRAS Project ID: 283281). Data sharing statement: Clinical Study Report, Protocol and data will be available on-line at ClinicalTrials.gov following publication.

## Abstract

There is evidence that some powered muscle stimulators can affect cardiac demand pacemakers, although devices vary enormously in terms of the intensity, frequency, and duration of stimuli. Regulatory authorities require that specialist medical opinion be obtained before using a stimulator on a patient with an implanted device. However, no explicit criteria are specified. This study aims to determine if there are any detectable interactions between a small, wearable stimulator which activates the muscle pumps of the leg by delivering a single momentary pulse at a frequency of 1Hz. 28 patients with pacemakers (single chamber, dual chamber, or bi-ventricular) were given stimulation. The pacemaker marker channel was recorded, and the electrogram examined to detect any sensed signal from the stimulator. In total, 2372 energy pulses were delivered by the stimulators. None of these were sensed by the pacemaker. This appears to show that there is no appreciable interaction between this type of stimulator and cardiac demand pacemakers

## Introduction

Powered muscle stimulators are covered by the United States Food and Drug Administration (FDA) Code of Federal Regulations CFR.890.585 (1). The code requires that the labelling for powered muscle stimulators includes the contraindication ‘Powered muscle stimulators should not be used on patients with cardiac demand pacemakers’. Further, IEC 60601-2-10:2012 clause 210.7.9.2.101d, requires that the instructions for use provide: “ Advice that a patient with an implanted electronic device (for example a cardiac pacemaker) should not be subjected to stimulation unless specialist medical opinion has first been obtained” . However, the criteria which the specialist should apply are not specified.

There is evidence that some powered muscle stimulators, and related devices can affect Cardiac Demand Pacemakers (2, 3). However, powered muscle stimulators used to elicit a continuous tetanic contraction of the muscle, for example for functional assistance, typically use sustained pulse trains at a frequency of at least 30Hz (4) to elicit muscle contractions. The electrodes for these devices may be placed at number of sites on the body depending on the application, including the legs, arms and torso, often with well-spaced electrodes that may be attached to different limbs, sending electrical impulses through the torso. Conversely, a systematic review (5) of the literature concerning the interaction of physical therapy (including electrical stimulation) and cardiac rhythm devices recognises that the risk of interference from electrical stimulation devices (such as TENS) is lower when the electrodes are placed further away from the cardiac rhythm device and when they operate at low frequencies (2Hz or less).

The geko^®^ range of devices (Firstkind ltd, Daresbury) are small, wearable neuromuscular stimulators which activate the venous muscle pumps of the leg (6, 7). They operate by delivering a single momentary (<1 ms) pulse at a frequency of 1Hz with electrodes placed topically, close together, at the knee.

The aim of this study was to determine if there are any detectable adverse interactions between the geko^®^ and pacemakers. The primary endpoint was percentage of pacemaker sensed geko^®^ pulses in a 30 second period. The secondary endpoints were the incidence of adverse events (AEs), incidence of serious AEs (SAEs), incidence of study treatment related AEs, and the incidence of investigational device related AEs.

## Materials and Methods

28 informed, consenting patients >18 years with pacemakers were recruited. To minimise risk, only patients who are not pacing dependent were selected for participation. Patients were allocated to one of three study arms based on the type of pacemaker implanted: single chamber, dual chamber, or bi-ventricular. Exclusion criteria were concurrent use of a neuro-modulation device, any medication deemed by the Investigator to interfere with the study treatment (e.g. systemic steroids), and participation in any other clinical study that might interfere.

The study had a multi-centre prospective linear design (see Figure 1).

**Figure 1.**
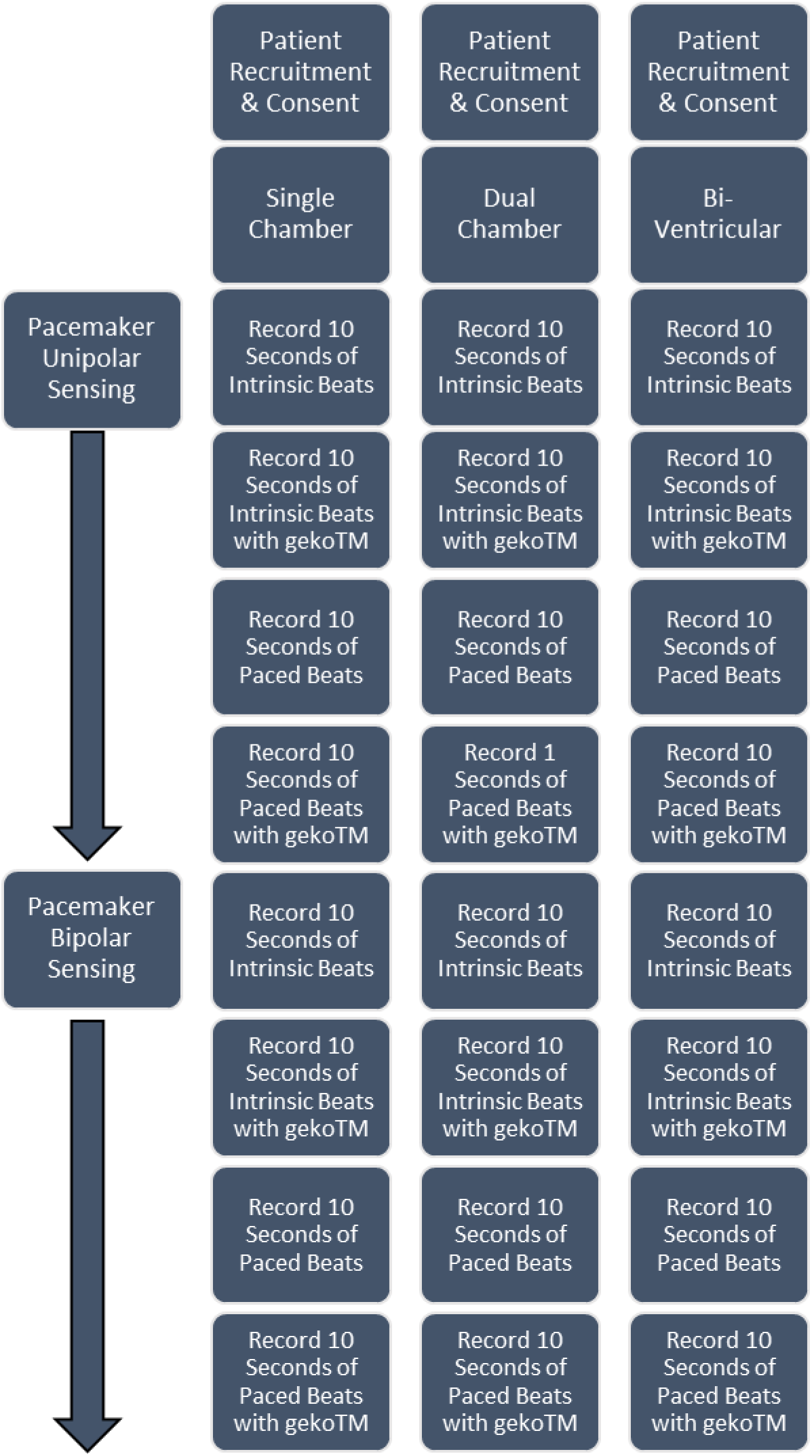
Schematic of study design.

Consented patients completed an initial baseline study. Here, they rested for a period of 10 minutes, then a sequence of 10 seconds of intrinsic beats were recorded and printed out on the pacemaker programmer using the internal pacemaker intracardiac electrogram (IEG).

Following the baseline, a geko^®^ was fitted to each leg, turned on and run for one minute and a further 10 second intracardiac electrogram recording made with the geko^®^ devices active. This process was repeated for paced beats without geko^®^ and finally paced beats with geko^®^. Additionally, the pacemaker was placed initially in unipolar sensing mode, then bipolar sensing mode (see Figure 1).

The geko^®^ devices were set at the maximum output level tolerated by the patient during the phases of the study where the device was active.

A pacemaker programmer was used to program the pacemaker and record the cardiac signal and marker channel. For the purposes of this study geko^®^ pulses sensed by a pacemaker are defined as *‘Sensed events on the pacemaker programmer marker channel which are not intrinsic cardiac signals and coincide with a geko*^®^ *pulse on the surface ECG’*.

A paper strip of the simultaneous marker channel, IEG and surface ECG was printed out from the pacemaker programmer for each recording’. It is this printout that was assessed by the investigator to calculate the percentage of sensed geko^®^ pulses sensed by the pacemaker (primary end point).

## Results

A summary of the patient demographics recorded in the study is detailed in Table 1.

**Table 1:** Patient population demographics.

|  | All Patients (N=28) |  | Single Chamber Patients (N=11) |  | Dual Chamber Patients (N=10) |  | Bi-Vent Patients (N=7) |  |
| --- | --- | --- | --- | --- | --- | --- | --- | --- |
|  | Mean | SE | Mean | SE | Mean | SE | Mean | SE |
| Height (m) | 1.669 | 0.019 | 1.681 | 0.024 | 1.669 | 0.012 | 1.649 | 0.018 |
| Weight(kg) | 81.829 | 5.243 | 81.836 | 5.130 | 84.300 | 6.446 | 78.286 | 4.079 |
| BMI | 29.297 | 1.827 | 29.071 | 1.885 | 30.183 | 2.302 | 28.390 | 1.028 |

Median age of pacemaker in the study was 3.68 years [IQR 1.85-5.94) Four major manufacturers were well represented in the cohort, see figure 2.

**Figure 2.**
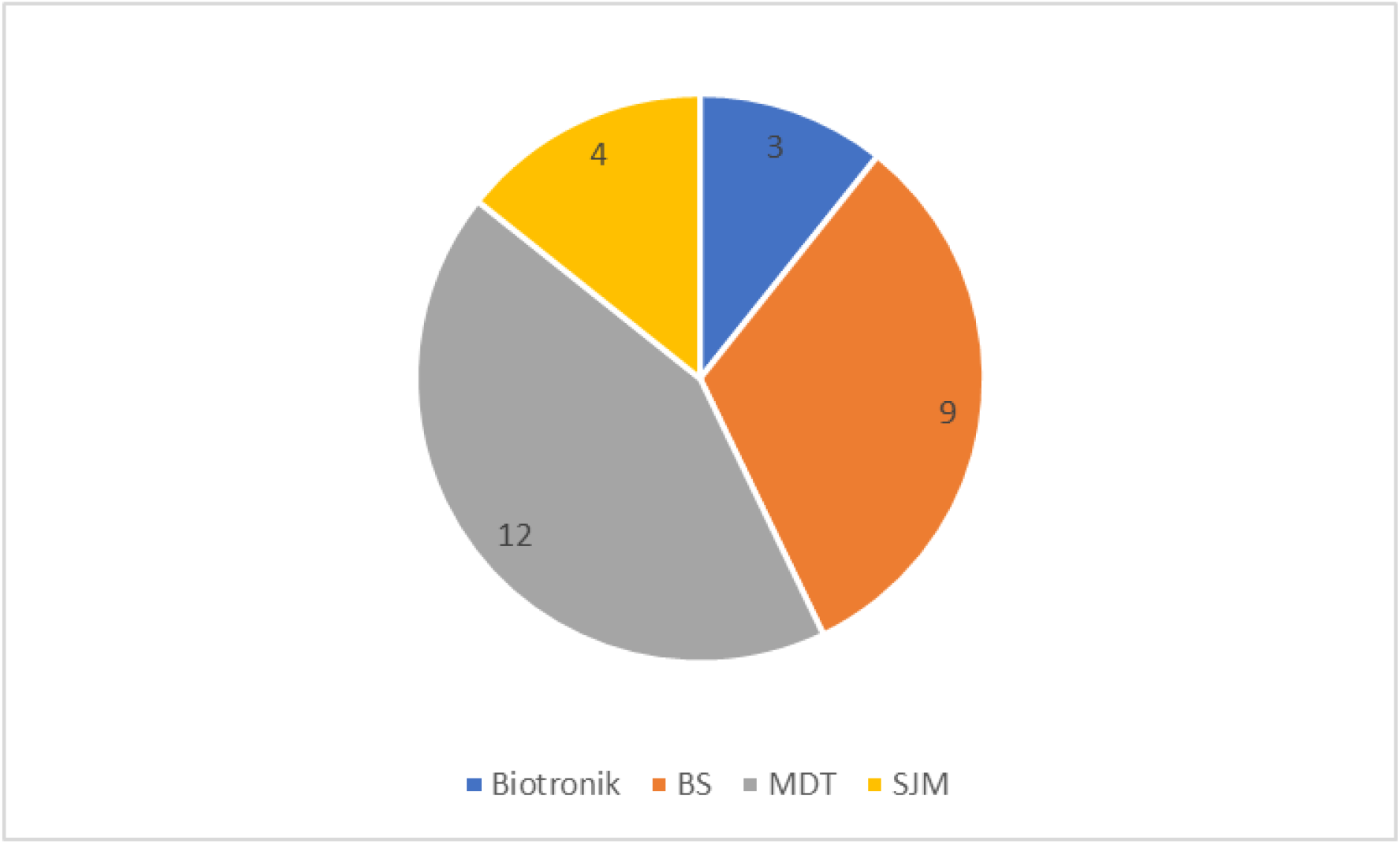
Pacemakers in study, by manufacturer

Every subject’s pacemaker performance was assessed by the investigator at baseline to ensure normal function. Test intracardiac electrogram strips were recorded and printed for each patient. These were repeated four times for each subject so that all sensing modalities (unipolar and bipolar) and pacing (paced cardiac rhythm) and non-pacing (intrinsic cardia rhythm) functionality could be assessed. In all cases stable and normal pacemaker function was observed, including lead impedance and pacing/sensing thresholds.

In total, 2372 energy pulses were delivered by geko^®^ devices to patients with implanted on-demand pacemakers. Pulses ranged across the energy output available for the device. Test intracardiac electrogram strips were recorded and printed for each patient. These were repeated four times for each subject so that all sensing modalities (unipolar and bipolar) and pacing (paced cardiac rhythm) and non-pacing (intrinsic cardia rhythm) functionality could be assessed. In all cases, the site investigator assessed the intracardiac pacemaker electrogram to determine if any energy pulses from the geko^®^ device had been sensed or recorded by the pacemakers; zero (0) geko^®^ pulses were sensed by pacemakers during this investigation.

A summary of the complete data set for all patients, pacemaker types and sensing configurations when the geko^®^ device is placed and delivering energy pulses to both legs is detailed in Table 2.

**Table 2:** Pacemaker-sensed geko^®^ pulses for all patients, pacemaker types and sensing configurations. Data from both legs.

| Pacemaker Sensing Configuration | geko<sup>™</sup> Pulses Delivered (N) | Mean geko<sup>™</sup> pulses per patient (N) | SE (mean geko<sup>™</sup> pulses per patient) | Mean Output Setting | SE (mean output Setting) | geko<sup>™</sup> pulse width at mean output setting (μs) | geko<sup>™</sup> power delivery at mean output setting (mA) | geko<sup>™</sup> pulses sensed by pacemaker (N) | Mean geko<sup>™</sup> pulses sensed by pacemaker (N) | SE (mean geko<sup>™</sup> pulses sensed by pacemaker) |
| --- | --- | --- | --- | --- | --- | --- | --- | --- | --- | --- |
| Combined data from all configurations. | 2372 | 21.179 | 0.194 | 8.286 | 0.168 | 280 | 38 | 0.000 | 0.000 | 0.000 |
| Intrinsic Cardiac Rhythm, Bipolar sensing | 604 | 21.571 | 0.382 | 8.286 | 0.338 | 280 | 38 | 0.000 | 0.000 | 0.000 |
| Intrinsic Cardiac Rhythm, Unipolar sensing | 580 | 20.714 | 0.335 | 8.286 | 0.338 | 280 | 38 | 0.000 | 0.000 | 0.000 |
| Paced Cardiac Rhythm, Bipolar sensing | 572 | 20.429 | 0.399 | 8.286 | 0.338 | 280 | 38 | 0.000 | 0.000 | 0.000 |
| Paced Cardiac Rhythm, Unipolar sensing | 616 | 22.000 | 0.431 | 8.286 | 0.338 | 280 | 38 | 0.000 | 0.000 | 0.000 |

No adverse events were recorded.

## Discussion

In this study, 2372 energy pulses were delivered by the trial device to 28 patients with variously single chamber, dual chamber, and bi-ventricular pacemakers. Of these 2372 pulses, 0 were detected by the pacemaker’s sensing function. This represents 0% of the sample, with an upper limit for the 95% confidence interval for sensed pulses of 0.25% as calculated by the Clopper-Pearson method (8).

Cardiac demand pacemakers operate by monitoring the P-wave of the heart’s ECG (9). The P-wave has a typical duration of 0.05 to 0.1s with an interval of about the same to the QRS-wave, the actual contractile pulse. To discriminate between the P-wave and any electromagnetic interference - such as 50-60Hz AC mains supply - cardiac demand pacemakers are fitted with low-pass filters to remove signals of this frequency and higher (10).

The geko^®^ pulse is a single square wave with a pulse width in the range 50-560µs, which have the same rise/fall dynamics as frequencies in the range 1785-20,000Hz, and thus may be expected to be removed by the pacemaker’s low-pass filter.

This concurs with Digby *et al* (5) systematic review of the literature concerning the interaction of physical therapy (including electrical stimulation) and cardiac rhythm devices, which ascribed a lower risk of interference from electrical stimulation devices when they operate at low frequencies (2Hz).

Badger *et al* (11) also conducted a systematic review, which was primarily focussed on Functional Electrical Stimulation (FES) and the potential for interaction with implantable cardioverter defibrillators ICDs). FES devices for drop-foot are similar to the geko^®^, in that, they apply electrical stimulation to the common peroneal nerve, albeit at a higher frequency. This group of authors concluded that FES devices for drop-foot could be considered safe when used on patients with pacemakers and ICDs. They also noted that there had been no case studies published concerning electromagnetic interference with pacemakers and ICDs when electrical stimulation was applied to the lower limb.

In conclusion, our analysis shows that there is no electro-magnetic interaction between the 1Hz monometric electromagnetic pulse delivered at the tibial site from the geko^®^ neuromuscular stimulator and a range of cardiac demand pacemakers. The geko^®^ device therefore appears to be safe to use in patients with implantable pacemakers.

## Data Availability

All data produced in the present study are available upon reasonable request to the authors

## Notes

### Competing Interest Statement

The authors have declared no competing interest.

### Clinical Trial

NCT04391257

### Funding Statement

The study was funded by Firstkind Ltd

### Author Declarations

Health Research Authority Wales REC 4 of the NHS gave ethical approval for this work

